# Impact of shaggy aorta on intraoperative cerebral embolism during carotid artery stenting

**DOI:** 10.1101/2022.05.02.22274007

**Authors:** Olesia O. Osipova, Savr V. Bugurov, Shoraan B. Saaya, Alexander A. Gostev, Alexey V. Cheban, Pavel V. Ignatenko, Andrey A. Karpenko

**Affiliations:** Center for Vascular and Hybrid Surgery, Meshalkin National Medical Research Center, Novosibirsk, Russian Federation

**Keywords:** carotid stenosis, transcranial Doppler ultrasonography, cerebral embolism, carotid artery stenting, shaggy aorta, stroke

## Abstract

**Background:** Careful selection of patients for carotid stenting is necessary. We suggest that patients with a shaggy aorta syndrome may have a higher risk of perioperative embolic complications.

**Methods:** The study is a subanalysis of the SIBERIA Trial. We included 72 patients undergoing transfemoral carotid artery stenting. All patients had an MRI DWI and a clinical neurological examination two days before and on the second and 30th days after the intervention.

**Results:** 46 patients had shaggy aorta syndrome. Intraoperative embolisms were recorded in 82.6% and 46.1% of patients with and without shaggy aorta syndrome, respectively (p=0.001). New asymptomatic ischemic brain lesions in the postoperative period occurred in 78.3% and in 26.9% of patients with and without shaggy aorta syndrome, respectively (p=0.000). 3 (6.5%) cases of stroke within 30 days after the procedure were observed only in patients with shaggy aorta syndrome. Shaggy aorta syndrome (OR 5.54 [1.83:16.7], p=0.001) and aortic arch ulceration (OR 6.67 [1.19: 37.3], p = 0.02) were independently associated with cerebral embolism. Shaggy aorta syndrome (OR 9.77 [3.14-30.37], p=0.000) and aortic arch ulceration (OR 12.9 [2.3: 72.8], p = 0.003) were independently associated with ipsilateral new asymptomatic ischemic brain lesions.

**Conclusions:** Shaggy aorta syndrome and aortic arch ulceration significantly increase the odds of intraoperative embolism and new asymptomatic ischemic brain lesions. Carotid endarterectomy or transcervical carotid stent should be selected in the presence of aortic ulceration.

## Introduction

Most RCTs (CREST, ACST-1, ACST-2) used strokes and deaths as endpoints to compare the efficacy of two carotid revascularization techniques (open and endovascular).^1^ The incidence of asymptomatic brain lesions after carotid stenting and carotid endarterectomy were not compared. Asymptomatic cerebral lesions can also have clinical significance and affect the patient’s cognitive status. ^2,3^ Despite the fact that carotid artery stenting (CAS) is a minimally invasive procedure, intraoperative cerebral embolism remains the main problem and can increase the risk of symptomatic and asymptomatic ischemic foci of the brain. It has been established that there is an association between microemboli during carotid revascularization and ipsilateral new brain lesions. ^4^

The use of new-generation anti-embolic protection systems and stents is aimed to reduce the number of embolisms, but the problem of cerebral artery microembolism has not yet been completely solved. Intravascular manipulation of catheters and guides in the aortic arch and carotid arteries can lead to the mobilization of atherosclerotic plaque fragments and cerebral microembolization.^5^ Manipulations can be particularly dangerous in patients with a shaggy aorta.

We suppose that the preoperative assessment of the aortic arch can help predict intraoperative microembolism and the risk of carotid artery stenting.

The aim of this study was to assess the relationship between the presence of a shaggy aorta, an intraoperative embolism and ipsilateral ischemic stroke or ipsilateral new symptomatic ischemic brain lesions on MRI DWI in patients with carotid stenting.

## Materials and methods

### Study design

The study is a subanalysis of the SIBERIA Trial (ClinicalTrials.gov Identifier: NCT03488199). The study design is shown in Figure 1. This study was conducted in accordance with the principles of the Declaration of Helsinki and Good Clinical Practice guidelines. The local ethics committee of the Meshalkin National Medical Research Center was approved. Written informed consent was obtained from all patients before enrollment.

**Fig. 1.**
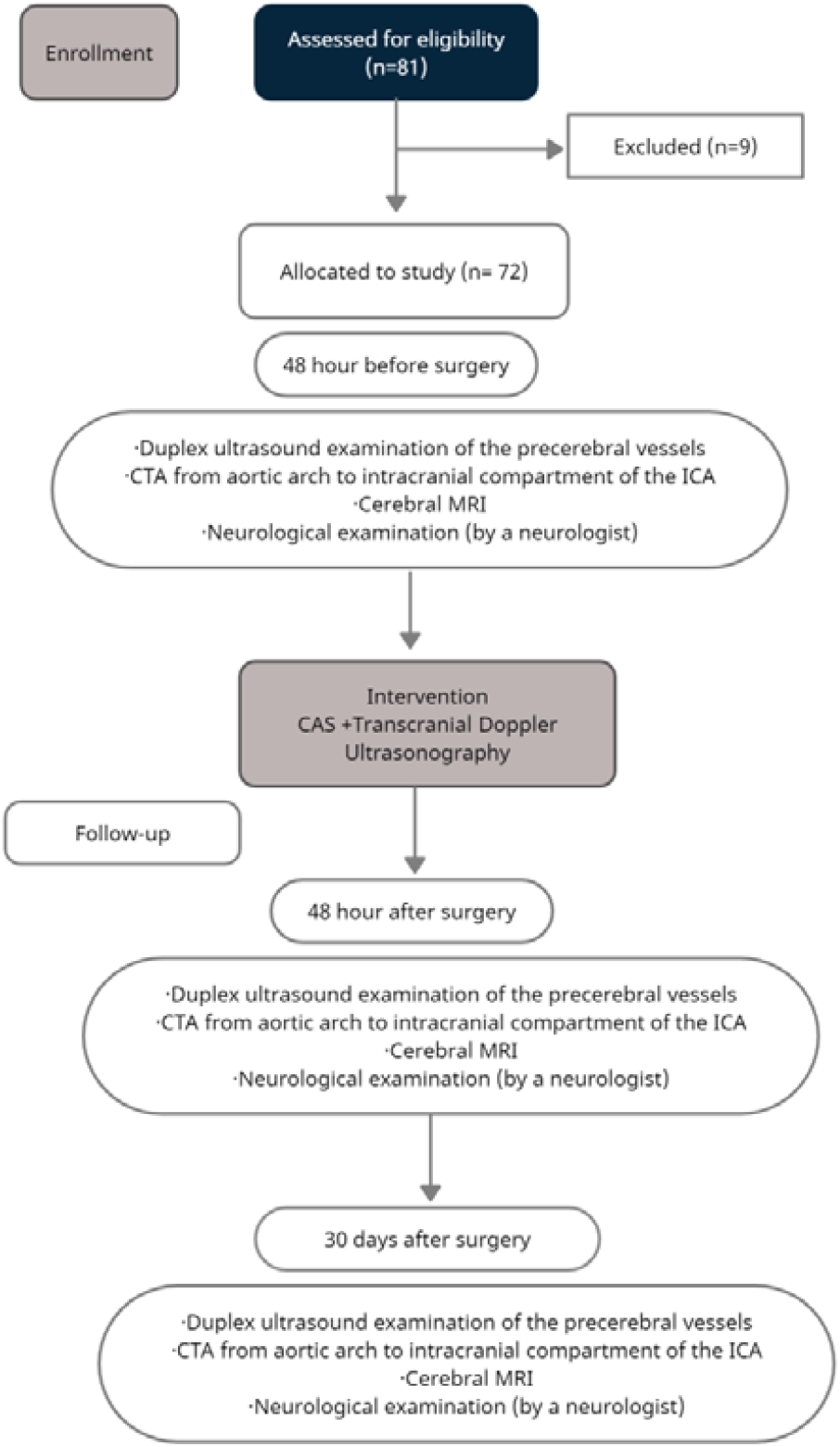
Flow Diagram of Study Design.

**Figure 2.**
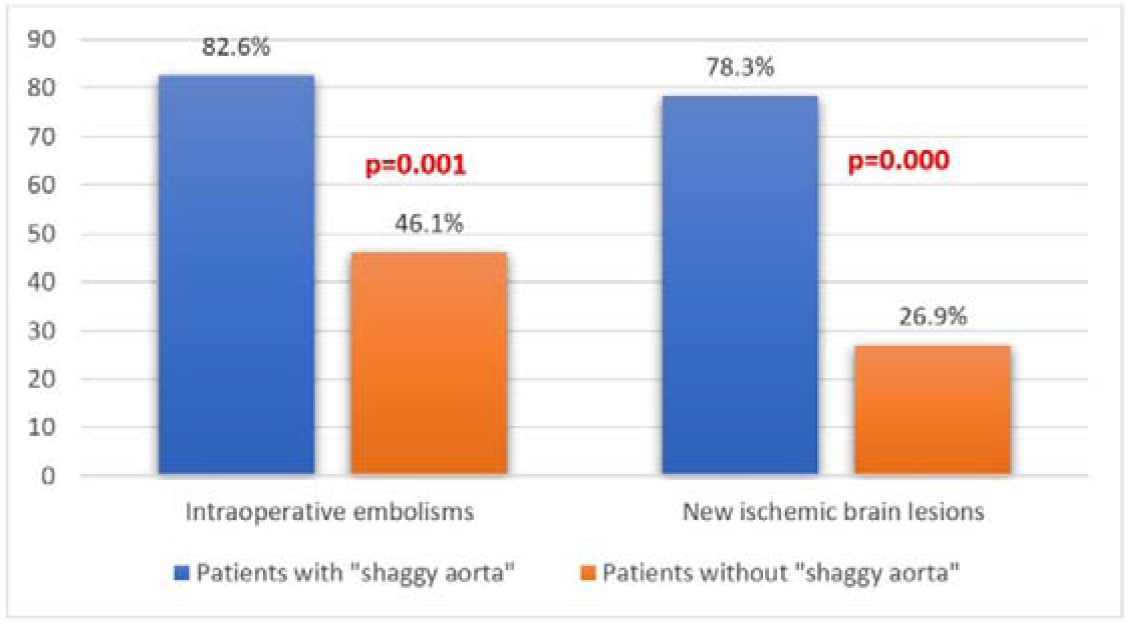
Frequency of Intraoperative Embolism and New Asymptomatic Ischemic Brain Lesions.

### Patient selection

The study was conducted from March 2019 to March 2020.

Inclusion criteria were: adult patient, informed consent, symptomatic ICA stenosis more than 60% and asymptomatic stenosis more than 70% according to NASCET criteria, blood flow velocity through the internal carotid artery based on Doppler ultrasound > 2 m/s, presence of a temporal window, high surgical risk (severe heart failure or severe pulmonary disease), consent to participate in research.

Exclusion criteria were: previous operation on the ipsilateral ICA, myocardial infarction less than 6 months ago, traumatic brain injury less than 3 months ago, hemorrhagic stroke within the last 3 months.

### Imaging

Color duplex ultrasonography of the precerebral vessels, computed tomographic angiography (CTA) of the aortic arches, carotid arteries and intracranial arteries, MRI DWI and clinical neurological examination were performed on all patients two days before, and on the second and 30th day after the intervention. Transcranial Dopplerography (TCD) was performed on all patients during CAS surgery to register embolism in the cerebral arteries.

#### Computed tomography angiography

CTA was performed at 64-320 slice CT scans. In pre-operational examination CTA was performed from aortic arch to intracranial compartment of the internal carotid artery. Voltage - 70-120 kV, pitch - individual, scan direction: cranio-caudal. Iodine contrast agent was used in all cases and administered at 4-5 mL/s intravenous (catheter size: 18-22 G). Contrast volume was calculated with the formula: contrast volume=speed contrast x scanning time. Saline was administered in the same volume as contrast at the same speed. The thickness reconstructions slice was 0.5 - 1 mm. The percentage of stenosis of the internal carotid artery was calculated according to the ECST/NASCET criteria.

Blinded data analysis was performed. Two independent radiologists were looking for signs of a shaggy aorta. Shaggy aorta syndrome was defined as having at least two of the following: the presence of a wall thickness of more than 4 mm, inner contour irregularities, ulceration, and an intraluminal thrombus.

#### Cerebral Magnetic Resonance Imaging

Brain MRI studies were performed before the procedure, 48 hours after CAS, and at 30 days using an Achieva 1.5-Tesla scanner (Phillips, The Netherlands). Core laboratory MRI imaging blinded data (sequential patient number-only) were transferred to the external core laboratory. Magnetic resonance offline analysis system (Syngo.via VB40, Siemens Healthcare, Germany) was used. Two independent radiologists (with over ten years of MRI cerebral image analysis experience), blinded to the treatment assignment, study timing, and the patient data, evaluated all MRI images qualitatively and quantitatively.

Reconstruction matrix was 256 or higher. The scans included DWI (diffusion-weighted imaging) with the automatic reconstruction of the measured diffusion coefficient in the scan protocol in addition to the usual modes of T1 and T2-weighted and fluid attenuation inversion recovery (FLAIR) imaging (T 1 SE, T 2 FRFSE, FLAIR iv FSE modes). The protocol for DWI sequences with an automatic reconstruction of the measured diffusion coefficient in the scan protocol was characterized by matrix 128×128 or higher, TE 84.2ms, TR 7000ms, voxel size-RL 0.92 mm, AP, FH, b values 0-100-1000). For T2 weighted sequences, typical TE was 86.2ms; TR 4240ms; voxel size RL 0.429 mm, AP, MS, FH), and (FLAIR) (TE 21ms; TR 9000ms; voxel size RL-0.83mm, AP, FH). The study was carried out in axial planes with a slice thickness of 5mm, with gap ≤ 15% of slice thickness.

Blind analysis of the data was carried out. Any disagreement between two core lab radiologists was resolved by consensus involving a third radiologist. The apparent diffusion coefficient (ADC) map (b values 0–600 mm/s2) was calculated. Acute ischemic lesions were defined as high signal intensity areas on DW images and low ADC values that were absent on baseline examination. Acute periprocedural lesions becoming permanent at 30 days were assessed on T2-weighted FLAIR images (same lesion location as the new DW-MRI lesion on post-procedural scan, FLAIR lesion absence on baseline scan). Lesion number, location, and size (volume in mm3) were analyzed and recorded.

##### Transcranial Doppler ultrasonography

In order to register embolism in the cerebral arteries, all patients underwent transcranial Doppler ultrasonography (TCD) continuously during CAS. In the course of the study, the apparatus “Angiodin-Universal” with a 2 / 2.66 (2.3) MHz sensor was used. We considered that we found the middle cerebral artery (MCA) in the presence of a pulse wave and its depth of 55-65 mm. The probe was positioned approximately 1 cm anterior to the external auditory canal and 1 cm above the zygomatic bone. By manipulating the sensor and changing the settings, the best view of the spectrogram of the MCA blood flow was selected.

Previously, 24 hours before the intervention, a preliminary search for MCA in patients was performed, and, as a consequence, the presence of a suitable temporal window. In the absence of one and the impossibility of monitoring the MCA, the patient was not included in the study.

Positioning and adjustment of the sensor was carried out before the intervention. Thus, in the case of stenting, the positioning of the transducer began before the puncture of the femoral artery. Monitoring ended after completion of the operation. During the monitoring, the device was tuned to suppress possible interference (pseudo-emboli). The analysis of the data obtained was carried out within 24 hours after the intervention by two experts.

###### Interventions

All surgical procedures were performed by specialists in vascular pathology and hybrid surgery. The procedures were performed under local anesthesia. All CAS procedures were performed through a transfemoral access with the Emboshield NAV^6^(tm) filter (Abbott Vascular, USA) as the distal embolic protection device. During the procedure, heparin was administered intravenously (5000 IU for patients weighing less than 70 kg and 7500 IU for patients weighing more than 70 kg). The following types of stents were used: open cell stent Acculink (Abbott Vascular, USA) and double-layer stent CGuard (Inspire MD, Israel, USA). The choice of the type of stent was left to the operator, depending on the specific case.

A distinction was made between 7 phases in order to assess the risk of each surgical step in the CAS procedure: carotid artery catheterization, installation of an embolic protection system, predilatation, stent placement, stent deployment, postdilation, removal of the embolic protection system.

### Medical therapy

All patients received aspirin (100-150 mg daily) indefinitely and clopidogrel (75 mg daily) at least 3 days before and 3 months after CAS. A loading dose of 300-mg clopidogrel was given during or after the intervention.

### Definitions

Any new focal neurologic deficit (caused by cerebral ischemia) lasting more than 24 hours was classified as ischemic stroke, while symptoms lasting less than 24 hours were classified as transient ischemic attacks.

### Endpoints

Clinical outcome measures were ipsilateral (to the procedure) cerebral ischemic stroke or transient ischemic attack (TIA) during or within 30 days of the procedure. We also assessed ipsilateral new asymptomatic ischemic brain lesions within 30 days of surgery.

The study endpoints were:

- association between the presence of intraoperative embolism and ipsilateral new asymptomatic ischemic brain lesions
- association between the presence of a shaggy aorta signs and intraoperative embolism
- association between the presence of a shaggy aorta signs and ipsilateral new asymptomatic ischemic brain lesions

### Statistical analysis

Statistical analyses included descriptive statistics with mean (±SD) and median (range) for continuous variables and number (percentage) for categorical variables. Logistic regression was used to assess the relationship between the presence of intraoperative embolism and ipsilateral new asymptomatic ischemic brain lesions, the presence of a shaggy aorta signs and intraoperative embolism or ipsilateral new asymptomatic ischemic brain lesions. P values<0.05 were considered significant. To assess the sensitivity and specificity of cerebral embolism by TCD for new ipsilateral ischemic lesions on MRI, a Roc curve was constructed.

All calculations were carried out using the statistical software SPSS 24.0 package (IBM, Armonk, NY, USA) and Statistica (StatSoft, USA).

## Results

### Participant

81 patients were screened for study entry, but 9 patients were excluded, due to exclusion criteria. The remaining 72 patients were included in the study and analyzed. All 72 patients completed a 30-day follow-up period.

### Baseline data

Shaggy aorta syndrome was detected in 46 patients. In 37 patients the aortic wall thickness was more than 4 mm, in 49 patients there was an irregularity of the inner contour of the aortic arch, in 35 patients there was aortic ulceration and in 6 patients an intraluminal thrombus. At the same time, in 3 patients from the general group, only inner contour irregularities were observed. We did not assign them to the shaggy aorta syndrome, since it was necessary to have at least two signs. Demographic data, cardiovascular risk factors, and preoperative characteristics of the carotid lesions are presented in Table 1. CGuard stents were implanted in 43 (59.7%) and Acculink in 29 (40.3%) cases.

**Table 1.** Baseline Characteristics and Characteristics of the Treated Lesions.

|  | **Patients with shaggy aorta**<br>**n=46** | **Patients without shaggy**<br>**aorta n=26** | **P** |
| --- | --- | --- | --- |
| Age, years | 64.1±6.6 | 62.5± 6.7 | 0.13 |
| Gender, male | 33 (71.7%) | 21 (80.7%) | 0.28 |
| Tobacco use | 17 (36.9%) | 10 (38.5%) | 0.54 |
| Hypertension | 46(100%) | 26 (100%) |  |
| Coronary artery disease | 45 (97.8%) | 26 (100%) | 0.63 |
| Heart failure | 45 (97.8%) | 26 (100%) | 0.63 |
| Renal insufficiency | 5 (10.8%) | 2 (7.7%) | 0.50 |
| Atrial fibrillation | 5 (10.9%) | 6 (23.1%) | 0.14 |
| Diabetes mellitus | 18 (39.1%) | 9 (34.6%) | 0.45 |
| TIA or stroke in the previous 6 month | 14 (30.4%) | 10 (38.5%) | 0.33 |
| Lesion of contralateral ICA | 13 (28.2%) | 7 (26.9%) | 0.56 |
| stenosis | 10 (21.7%) | 4 (15.4%) |  |
| occlusion | 3 (6.5%) | 3 (11.5%) |  |

For continuous variables, the data is presented as mean ± standard deviation. For binary variables, the data is presented as weighted count (percentage).
Abbreviations: TIA: transient ischemic attack; ICA: internal carotid arteries.

### Main results

Cerebral microemboli were detected in all 38 (52.8%) patients during the CAS procedures. The median number of emboli in each patient during the procedure was 11 (range, 9 to 15).

Brain embolisms were registered during the introduction of catheters in 38 patients (52.8%), embolic protection system placing in 22 patients (30.55%), during predilation and stent placement in 5 (6.95%) patients, stent implantation in 3 (4.2%), postdilation in 7 (9.7%) cases, and removal of the protection filter in 1 patient (1.4%).

Intraoperative embolisms were recorded in 82.6% and 46.1% of patients with and without shaggy aorta syndrome, respectively (p=0.001). Ipsilateral new asymptomatic ischemic brain lesions in the postoperative period occurred in 78.3% amd in 26.9% of patients with and without shaggy aorta syndrome, respectively (p=0.000) (fig.2). 3 (6.5%) cases of stroke within 30 days after the procedure were observed only in patients with shaggy aorta syndrome. There were no cases of transient ischemic attack.

Among 38 general group patients with cerebral emboli by TCD, 30 patients had ipsilateral new asymptomatic ischemic brain lesions by MRI and 8 patients did not have them. Among 34 patients who did not have ipsilateral new ischemic lesions on MRI, 31 patients had no intraoperatively recorded episodes of microembolism and 3 patients had. The sensitivity and specificity of TCD was 78.9% and 91.1%, respectively. A roc curve was constructed to determine the sensitivity and specificity of transcranial Doppler for ipsilateral new asymptomatic ischemic brain lesions. (fig. 3). A categorical variable was used as a test variable (presence or absence of cerebral embolism according to TCD). The area under the curve was 0.85.

**Figure 3.**
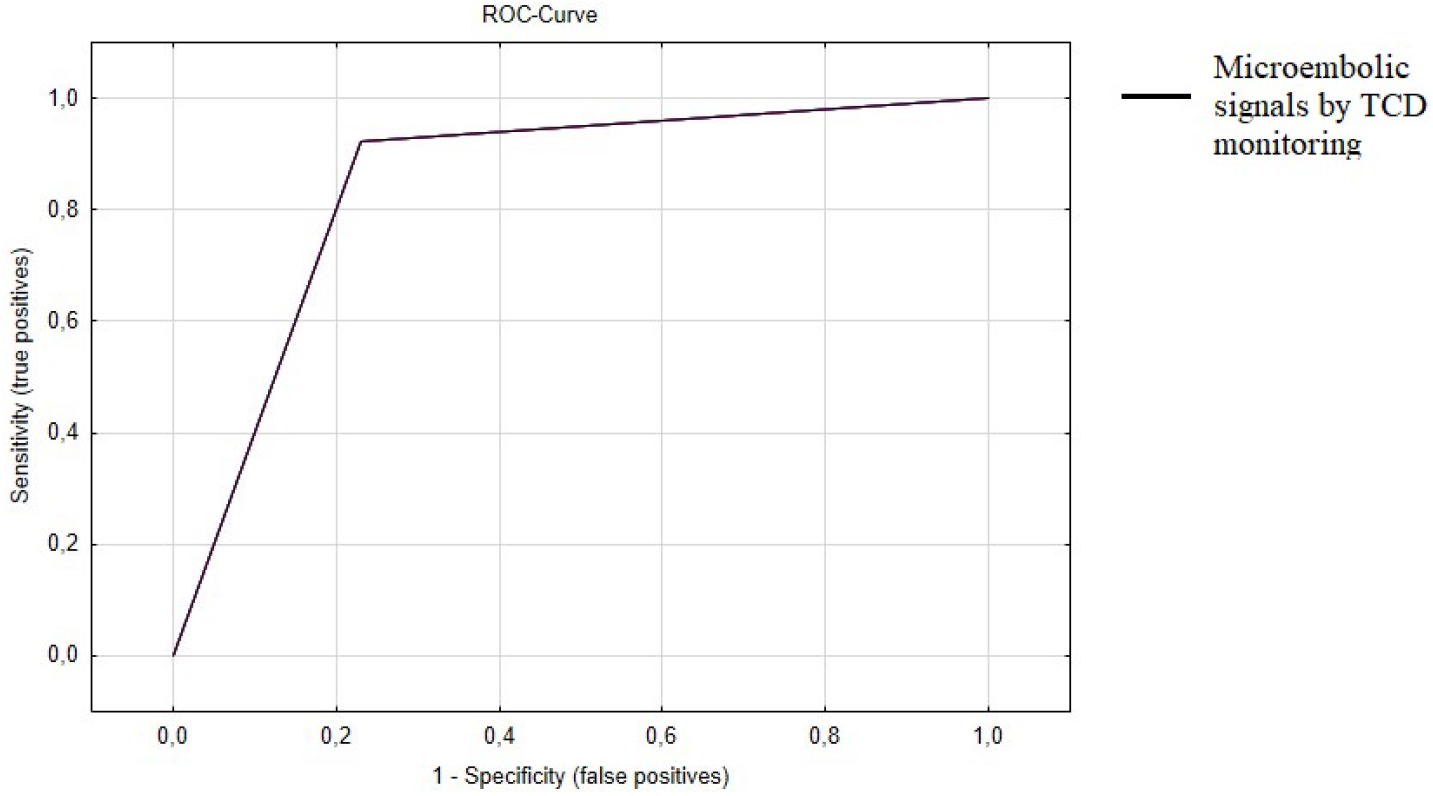
Receiver Operating Characteristic (ROC) Curve Plotted for Predicting Risk of Ipsilateral New Asymptomatic Ischemic Brain Lesions.

Logistic regression analysis showed a significant association between new ipsilateral ischemic brain lesions on MRI DWI and cerebral embolism by TCD (OR 25.68 [7.16: 92.06], p = 0.000).

The shaggy aorta syndrome was associated with cerebral embolism by TCD (OR 5.54 [1.83:16.7], p = 0.001) and ipsilateral new asymptomatic ischemic brain lesions (OR 9.77 [3.14: 30.37], p = 0.000 (Table 2, 3) The aortic arch ulceration was associated with cerebral embolism by TCD (OR 6.67 [1.19: 37.3], p = 0.02) and ipsilateral new asymptomatic ischemic brain lesions (OR 12.9 [2.3: 72.8], p = 0.003 (Table 2, 3)

**Table 2.** Factors Associated with Cerebral Embolism by TCD.

|  | Odds ratio | CI lower limit | CI upper limit | P-value |
| --- | --- | --- | --- | --- |
| Age, years | 0.98 | 0.88 | 1.09 | 0.75 |
| Sex, Female vs. Male | 0.86 | 0.01 | 0.67 | 0.017 |
| Type of stent, CGuard vs. Acculink | 0.47 | 0.1 | 2.09 | 0.31 |
| Shaggy aorta syndrome | 5.54 | 1.83 | 16.71 | 0.001 |
| Plaque thickness of > 4 mm | 1.94 | 0.32 | 11.5 | 0.45 |
| Inner contour irregularities | 1.27 | 0.29 | 5.48 | 0.73 |
| Ulceration | 6.67 | 1.19 | 37.3 | 0.02 |

Abbreviations: CI: confidence interval

**Table 3.** Factors Associated with New Asymptomatic Ischemic Cerebral Lesions.

|  | Odds ratio | CI lower limit | CI upper limit | P-value |
| --- | --- | --- | --- | --- |
| Age, years | 1.01 | 0.9 | 1.13 | 0.84 |
| Sex, Female vs. Male | 0.17 | 0.029 | 0.99 | 0.045 |
| Type of stent, CGuard vs. Acculink | 0.48 | 0.1 | 2.34 | 0.35 |
| Shaggy aorta syndrome | 9.77 | 3.14 | 30.37 | 0.000 |
| Plaque thickness of > 4 mm | 5.09 | 0.78 | 33.03 | 0.08 |
| Inner contour irregularities | 2.17 | 0.54 | 8.66 | 0.26 |
| Ulceration | 12.88 | 2.27 | 72.84 | 0.003 |

Abbreviations: CI: confidence interval

There were no statistical associations between stent type and cerebral intraoperative embolism and ipsilateral new asymptomatic ischemic brain lesions (Table 2, 3).

## Discussion

In this study, it was shown that shaggy aorta syndrome and aorta ulceration increases the odds of new ipsilateral ischemic cerebral lesions in patients undergoing stenting of the carotid arteries. In addition, the greatest number of embolisms was recorded at the stages of the catheters introduction and embolic protection system placing. This can be explained by the fact that the manipulation of catheters in the area of aortic arch and the brachiocephalic arteries carries a high risk of damaging the atherosclerotic plaque, especially if the plaque initially has an unstable surface.

Based on our data, we can conclude that preoperative detection of signs of a shaggy aorta is necessary to assess the risk of carotid stenting. The term "shaggy aorta” describes the severe aortic surface degeneration, extremely friable, and likely to cause atheroembolism. At the moment, the term shaggy aorta is quite variable, and in our opinion, the most appropriate definition is presented in the work of Kwon H. et all. ^6^ Shaggy aorta was defined as a diffuse, irregularly shaped atherosclerotic change involving > 75% of the length of the aorta from the aortic arch to the visceral segment with an atheromatous plaque thickness of > 4 mm. In our study, both episodes of intraoperative embolism and new ischemic cerebral lesions were associated with shaggy aorta syndrome and aortic arch ulceretion.

If signs of a shaggy aorta are found, it may be worth abandoning stenting in favor of carotid endarterectomy or performing a transcervical stenting procedure. Changing the standard transfemoral access to the transcervical one may reduce the number of microembolic complications during CAS, since this excludes navigation by catheters and guide wires through the aorta and carotid arteries. The ROADSTER study showed that the 30-day stroke risk in 144 high-risk patients who underwent transcervical CAS with retrograde blood flow was 1.4%. ^7^

Our study confirmed the association between new ipsilateral ischemic cerebral lesions and intraoperative embolic signals. The values of the areas under the Rock curves, which tended to 1, indicates high predictive ability. Previous studies have also shown an association of solid and gaseous microemboli with periprocedural ipsilateral ischemic strokes or new diffusely weighted cerebral MRI lesions.^8–10^ There has been a proven correlation between microembolic signals on TCD and stroke / TIA.^11^

Half of the patients had new ischemic cerebrallesions on MRI DWI. Despite the fact that most cerebral ischemic lesions remained asymptomatic, they can have clinical significance in the long-term period.

Microembolisms during CE and CA surgery can lead not only to the development of a stroke, but also to decrease in cognitive functions. Cognitive disorder was not assessed in our study, but the Laza study, involving 32 patients, showed that microembolic signals on the TCD during the CAS procedure correlated with a decrease in cognitive functions during 1-year of follow-up. ^12^

It should be added that in our study, the open-cell stent did not become a predictor of microembolic events during CAS. There are quite conflicting data on the ability of stents of various designs to prevent protrusion of atherosclerotic plaque through the stent cells. ^13–18^

## Study limitations

This study has potential limitations. The study had a relatively small sample size. In the future, a comparative study should be carried out in patients undergoing open surgery, transfemoral and transcervical carotid stenting.

## Conclusion

Before planning a carotid revascularization method, it is necessary to evaluate the aortic arch for signs of shaggy aorta syndrome. The presence of shaggy aorta syndrome and aortic ulceration is associated with intraoperative cerebral embolism and ipsilateral new asymptomatic ischemic brain lesions. Considering that introduction of catheters and embolic protection system placing led to most number intraoperative embolisms, if there are CT signs of a shaggy aorta according, the stenting procedure should be abandoned in favor of an open operation or transcervical access should be chosen.

## Data Availability

All data produced in the present work are contained in the manuscript

## Acknowledgments

the investigators thank Bergen T.A., Bobrikova E.E. for the analysis of computed tomography angiography and cerebral magnetic resonance imaging

## Abbreviations

CAS: carotid artery stenting
CTA: computed tomography angiography
TIA: transient ischemic attack
ICA: the internal carotid arteries
MCA: the middle cerebral artery
TCD: transcranial Doppler sonography

